# Maternally-derived antibody titer dynamics and risk of hospitalised infant dengue disease

**DOI:** 10.1101/2022.11.18.22282500

**Authors:** Megan O’Driscoll, Darunee Buddhari, Angkana Huang, Adam Waikman, Surachai Kaewhirun, Sopon Iamsirithaworn, Direk Khampaen, Aaron Farmer, Stefan Fernandez, Isabel Rodriguez-Barraquer, Anon Srikiatkhachorn, Stephen Thomas, Timothy Endy, Alan L. Rothman, Kathryn Anderson, Derek A.T. Cummings, Henrik Salje

**Affiliations:** Department of Genetics, University of Cambridge, UK; Department of Virology, Armed Forces Research Institute of Medical Sciences, Bangkok, Thailand; Department of Microbiology and Immunology, State University of New York Upstate Medical University, New York, USA; Department of Disease Control, Ministry of Public Health, Tiwanond, Nonthaburi, Thailand; University of California, San Francisco, USA; Department of Cell and Molecular Biology, Institute for Immunology and Informatics, University of Rhode Island, Providence, RI, USA; Faculty of Medicine, King Mongkut’s Institute of Technology Ladkrabang, Thailand; Department of Medicine, State University of New York Upstate Medical University, New York, USA; Coalition for Epidemic Preparedness Innovations, USA; Department of Biology, University of Florida, USA

## Abstract

Dengue virus (DENV) immunity is complex. Maternally-derived DENV antibodies initially provide protection against infection, however, as antibodies decay they can enhance disease severity upon infection. Quantifying antibody titers that are associated with disease risk is complicated by their dynamic nature and imperfect measurement processes. It also remains unknown whether long-term trends in birth rates, population-level infection risks, and maternal ages have altered immune profiles in child-bearing women, leading to shifts in age-specific infant disease risks. Here, we analyse DENV antibody data from two infant cohorts (N=165 infants with 665 blood draws) and 40 years of infant dengue hospitalisation data from Thailand. We use mathematical models to reconstruct maternally-derived antibody dynamics and estimate hospitalisation risk by titer. We find the relative risk of dengue hospitalisation ranges from 0.13 (0.00-0.89) in 1 month olds to 3.52 (3.25-3.79) in 8 month olds, compared to the risk in 12 month olds. We estimate the highest risk of infant dengue hospitalisation occurs at PRNT_50_ titers of 6.0 (5.7-6.6). Our inferred titer-related risk estimates are consistent with previously identified titer-based correlates of severe disease among older individuals experiencing secondary DENV infections, suggesting a common mechanism of risk enhancement from pre-existing antibodies. Finally, we describe how the shifting epidemiology of dengue in Thailand, combined with declining birth rates, have decreased the absolute risk of infant dengue disease by 64% over a 40 year period while having minimal impact on the mean age of infant hospital-attended dengue disease.

## Introduction

Dengue virus (DENV) is a mosquito-borne flavivirus with four serotypes (DENV1-4). As in many tropical regions, DENV is hyper-endemic throughout South-East Asia. While primary DENV infections are typically mild or asymptomatic, secondary infections are associated with an increased risk of severe disease. This increased risk is thought to be mediated by a mechanism known as antibody-dependent enhancement (ADE), whereby pre-existing heterologous antibodies bind but do not neutralise infecting virions (1–3). Binding of non-neutralising antibodies to the infecting serotype is believed to facilitate virus entry into Fc-receptor-bearing cells and subsequently, increase viral burden in the host (1, 4, 5). Antibody concentration is therefore likely a key determinant of ADE and gives rise to the idea of a titer-related “window” of risk, where titer levels are low enough that they do not effectively neutralise virions but high enough that they enhance heterologous infections (2, 3, 6).

ADE is also believed to occur in young infants when experiencing their primary infection if maternally-derived anti-DENV antibodies are present (7–11). The placental transfer of IgG antibodies from mother to infant is an important mechanism for protecting young infants from infectious pathogens while the neonatal immune system is still developing (Ciobanu et al., 2020). These transplacentally-acquired antibodies decay within the first year of life, initially providing protection against infection, and subsequently waning over time to sub-neutralising levels that have the potential to mediate ADE (illustrated in Figure S1). As antibody levels decay further, the ability to enhance the severity of heterologous infections is thought to be lost, though infants remain at risk of DENV infection. Peaks in reported dengue hospitalisations among infants less than 1 year of age are consistently observed in DENV-endemic countries and are widely attributed to ADE (7, 11, 12). However, the specific concentrations of maternally-derived DENV antibodies that may place infants at increased risk of severe disease have not been well characterised.

Quantifying the dynamics of maternally-derived antibodies and associated risks of severe dengue disease requires carefully designed cohort studies with regular blood draws. However, very large cohort sizes are required to observe a sufficient number of severe dengue outcomes with which to directly investigate titer-associated risks, often making such studies infeasible. A further complication to our understanding of the relationship between pre-existing DENV antibodies and risk of severe disease upon infection is that any antibody concentration-associated window of risk may be masked by lower limits of assay detection (typically a dilution of 1:10). For example, a previous study in Vietnam found that DENV-neutralising antibodies declined to undetectable levels in >90% of infants by 6 months of age (13). In addition, many assays measure antibody concentration as endpoint titers defined as the reciprocal of the highest sample dilution that gives a measurable effect. Typically conducted using 2-fold serial dilutions, this discretized measurement process obscures the true underlying continuous concentration values, adding to the challenge of disentangling antibody titer dynamics and associated risks.

Mathematical modelling can aid our understanding of these unobserved mechanisms. By accounting for assay characteristics such as the lower limit of detection and discretized measurements, the underlying temporal trends in antibody titers can be inferred. Given the immunological vulnerability of infants in their first year of life, there is a clear need to characterise the timing and duration of periods when they are at greatest risk of severe outcomes, as well as to understand the mechanisms that mediate their risk. In this study we use data from two longitudinal cohort studies conducted in Thailand which recruited infants at birth and measured DENV antibody levels at regular follow up visits. We combine these data with the age distribution of infant dengue cases reported in local hospitals over a forty year time period to probabilistically infer the population titer distributions associated with the age of enhanced risk of dengue hospitalisation in infants. Finally, we use data on the changing profiles of fertility rates, mean age of childbearing and DENV transmission intensity in Thailand over the past 40 years to explore how these trends may influence infant DENV risk profiles.

## Results

We used longitudinal serological data from cohorts in Bangkok (2000-2001, N=123 infants) and Kamphaeng Phet (2015-2021, N=45 infants), Thailand. We excluded all infants that had evidence of a DENV infection during the first year of life, as defined by increases in mean DENV titers at sequential blood draws. Geometric mean cord blood titers at time of birth across the four serotypes in the Bangkok cohort were 303.8 (95%CI: 254.2-363.1) as measured by plaque reduction neutralisation test (PRNT_50_) and 32.9 (95%CI: 29.7-36.4) as measured by hemagglutination inhibition (HI) assay (Figure 1A-B). Mean PRNT_50_ titers fell to 47.2 (95%CI: 41.1-54.2) by three months and to 9.6 (95%CI: 8.8-10.6) by six months, by which time 62.7% of PRNT_50_ titers were below the limit of detection. Mean HI titers fell to 13.0 (95%CI: 12.0-14.1) at 3 months and to 6.8 (95%CI: 6.4-7.2) at 6 months. By 12 months, 98.8% of PRNT_50_ and 100% of HI measurements were below the limit of detection. A similar pattern was observed in the Kamphaeng Phet cohort, with geometric mean cord blood HI titers of 99.3 (95%CI: 81.5-121.1), falling to a mean of 46.8 (95%CI: 38.4-57.0) by 3 months and 10.0 (95%CI: 8.3-12.1) by six months (Figure 1C). At six months, 57.3% of titers were below the limit of detection, rising to 93% at 12 months. Antibody titers in mothers’ sera were strongly correlated with cord blood titers (Pearson correlation coefficient, R=0.94 in Bangkok for PRNT_50_, and 0.87 for HI in Bangkok) (Figure S2). Cord blood titers were also strongly correlated with infant serum titers at ages <6 months, though the correlation decreased with increasing infant age (Figure S2).

**Figure 1.**
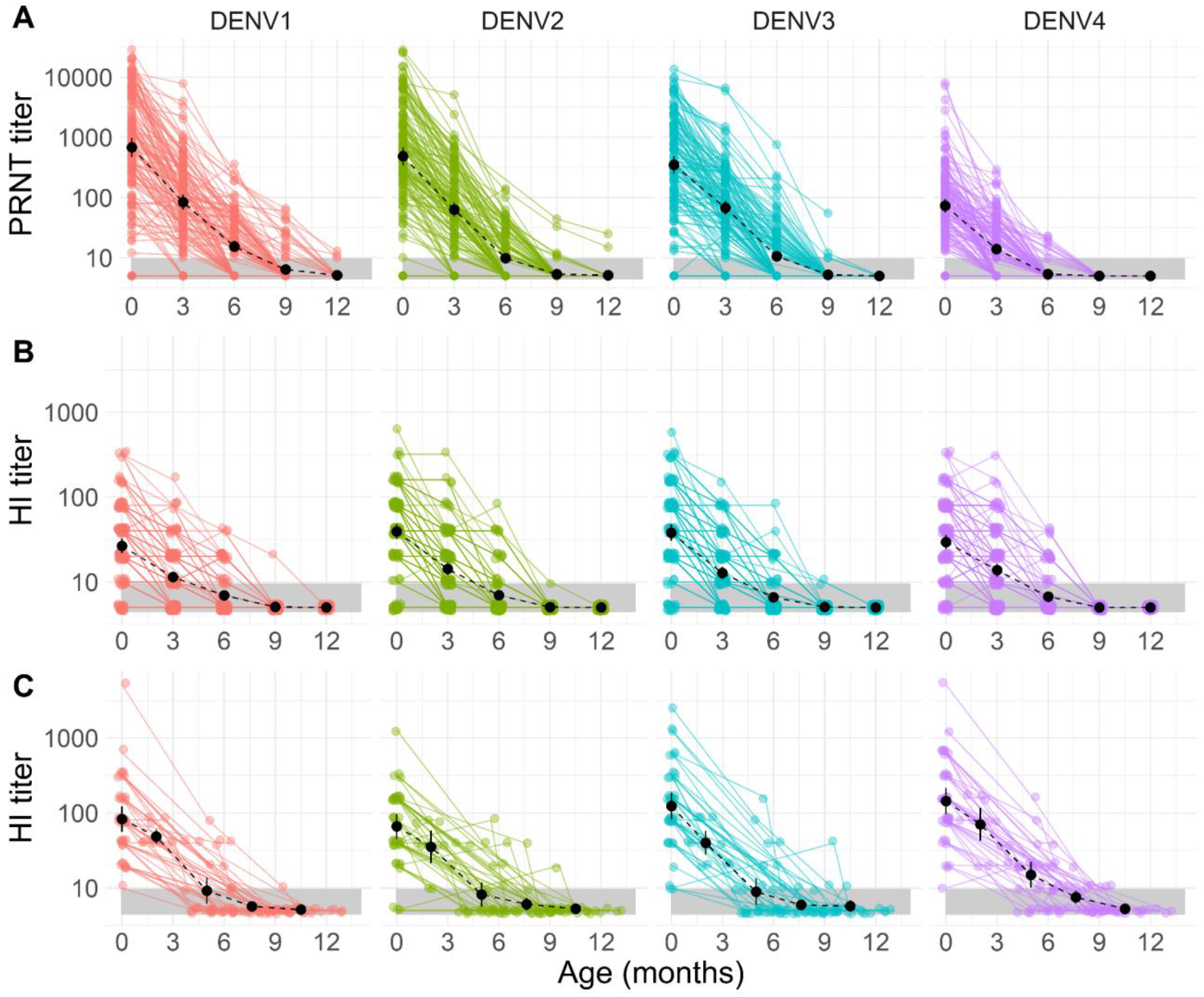
DENV antibody titer trajectories among infants aged 0-13 months by serotype. Observed titers by age observed in the Bangkok cohort as measured by the PRNT and HI assays (Panels A and B, respectively) and HI titers from the Kamphaeng Phet cohort (Panel C). Titer values are shown on a log-linear scale in all panels. Coloured points and lines show the observed titers and trajectories of individual infants. Black points and lines represent the geometric mean titer and 95% confidence intervals across infants at each age group. The black dashed line shows the trajectory of geometric mean titers across infants by age. The grey shaded area highlights the lower limit of detection (titers <10) of the HI and PRNT assays.

We developed a mathematical model to reconstruct maternally-derived antibody titers over the first 12 months of life in the two cohorts and explored a range of model parameterizations. The best fitting model, as assessed by deviance information criterion (DIC), assumed a time-constant rate of antibody decay and estimated separate decay rates for each assay and cohort (Table S1). We estimated a median half-life of 28.5 days (95% credible interval (CrI): 27.6-29.4) for PRNT_50_ titers in the Bangkok cohort, a half life of 47.1 days (95%CrI: 44.9-49.3) for HI titers in the same cohort and a half-life of 38.6 days (95%CrI: 35.6-41.5) for HI titers in the Kamphaeng Phet cohort. For the remainder of the results we focus on estimates from this best performing model. The estimated population-level maternally-derived antibody dynamics are shown by the solid green lines in Figure 2A-C. We compare these estimates to the observed data at each time point, accounting for the discretized measurement process of the assay (HI) and the lower limits of assay detection, shown by the black (observed) and green (estimated) points in Figure 2A-C. We find significant underlying differences in the antibody titers across serotype, with no clear trends by cohort and assay (Figure 2D).

**Figure 2.**
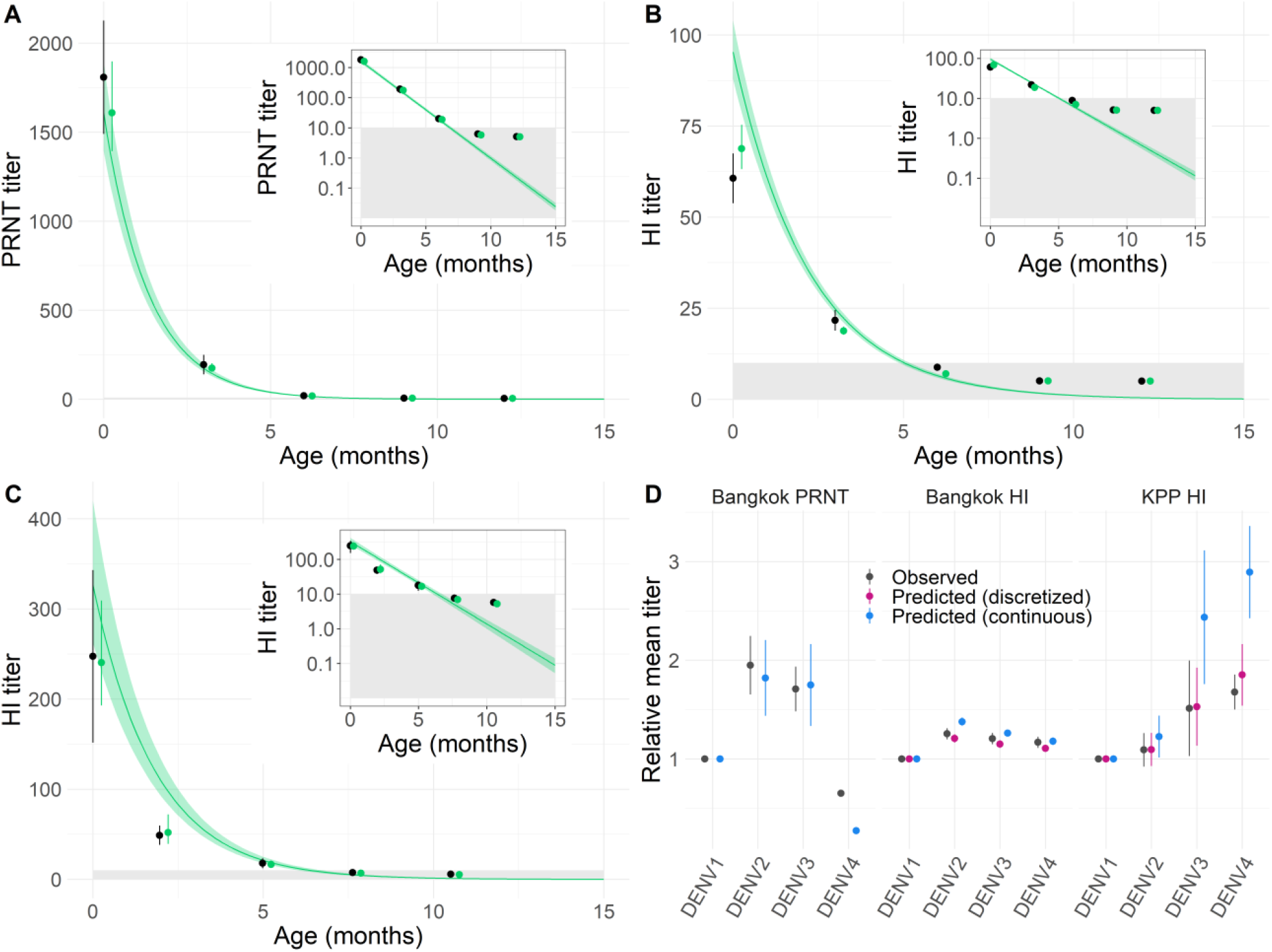
Observed and predicted population mean DENV antibody titers by cohort and assay. The green lines and ribbons show the median and 95% credible intervals of the estimated “true” population mean maternally-derived antibody titer from birth to 15 months (Panels A-C). Fits to data from the Bangkok cohort for PRNT and HI assays are shown in Panels A and B respectively, and from the Kamphaeng Phet cohort in Panel C. The black dots represent the mean and 95% confidence interval of the observed (discretized) titers and the green points represent the mean and 95% confidence intervals of the estimated underlying “true” titers on a discrete scale mimicking the discretized measurement process and lower limit of detection truncation. The insets in Panels A-C show model fits using a log-linear scale for antibody titers. The mean titer of each serotype, relative to DENV1, by cohort and assay are shown in Panel D where points and lines represent the median and 95% credible intervals.

A total of 1,422 infant dengue cases ≤12 months of age were reported at Queen Sirikit National Institute of Child Health (QSNICH) in Bangkok between 1974 and 2014, and 165 cases at Kamphaeng Phet (KPP) hospital between 1994 and 2019 (Figure S3). The mean age of reported infant (≤12 months) dengue hospitalisations was 7.3 (95%CI: 7.2-7.5) months in QSNICH and 7.9 (7.5-8.3) months in KPP hospital. The age distribution of hospitalised dengue cases for all ages and for infants only, joint across Bangkok and Kamphaeng Phet, are shown in Figure 3A-B. Across the two hospitals we estimate a mean age of 7.4 months (95%CI: 7.3-7.5) among infant dengue cases ≤12 months of age, with very little variability in the mean age of infant cases by year since 1974 (Figure 3C, p-value for a trend from linear regression of 0.53). Using the risk of infant hospitalisation in 12 month olds as a reference point and assuming a constant risk of exposure for all infants ≤12 months, we estimate the relative risk (RR) of infant hospitalisation with dengue to range from 0.1 (95%CI: 0-0.6) in 1 month old infants to 3.5 (95%CI: 3.3-3.8) in 8 month old infants (Table 1). There was no significant difference in the mean age of infant dengue cases by sex, or by month of hospitalisation (Figure S4). The mean age of infant DENV-4 cases was lower than that of other serotypes (Figure S4).

**Table 1.** Relative risk (RR) of infant dengue hospitalisation by month of age, relative to the risk in 12 month old infants.

| **Age (months)** | **Observed RR (95% CI)** | **Predicted RR (95% CI)** |
| --- | --- | --- |
| 1 | 0.13 (0.00-0.89) | 0.18 (0.00-0.85) |
| 2 | 0.33 (0.00-0.82) | 0.45 (0.00-0.92) |
| 3 | 0.71 (0.34-1.09) | 0.91 (0.53-1.28) |
| 4 | 1.62 (1.31-1.92) | 1.62 (1.29-1.95) |
| 5 | 2.22 (1.93-2.51) | 2.49 (2.19-2.79) |
| 6 | 3.22 (2.95-3.50) | 3.33 (3.04-3.62) |
| 7 | 3.05 (2.77-3.32) | 3.85 (3.57-4.14) |
| 8 | 3.52 (3.25-3.79) | 3.89 (3.60-4.18) |
| 9 | 2.92 (2.64-3.20) | 3.38 (3.09-3.67) |
| 10 | 1.95 (1.66-2.25) | 2.56 (2.26-2.87) |
| 11 | 1.56 (1.25-1.86) | 1.71 (1.38-2.03) |
| 12 | 1.00 | 1.00 |

**Figure 3.**
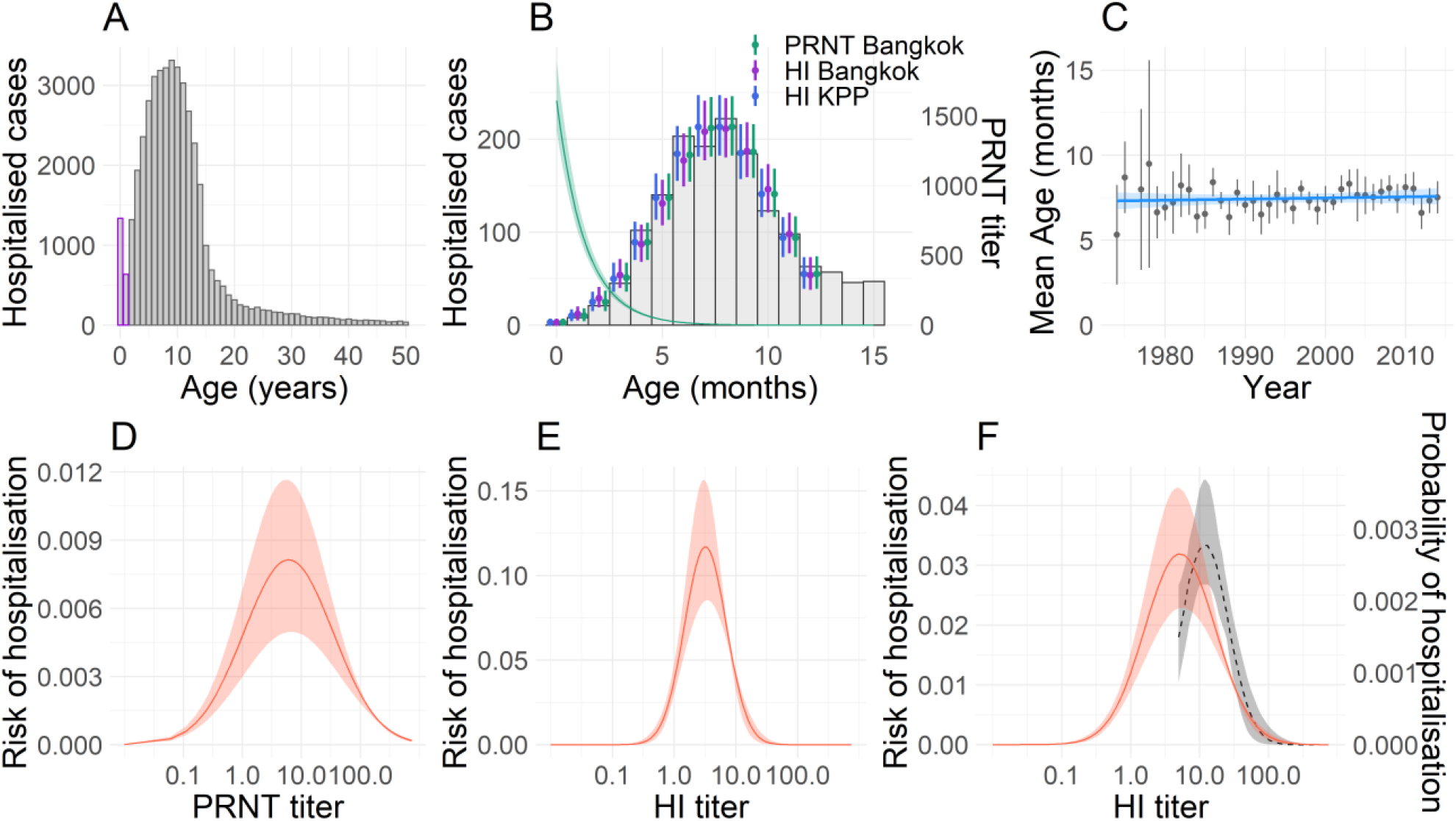
Age-specific risks of severe infant dengue disease and associated antibody titers. **(A)** The age distribution of all hospitalised dengue cases in QSNICH and Kamphaeng Phet hospitals aged between 0 and 50 years are shown by the grey bars. Infant cases aged 0-24 months are shown by the bars outlined in purple. **(B)** The age distribution of infant hospitalised dengue cases aged 0-15 months are shown by the grey bars. Green line and ribbon show the median and 95% credible interval of the expected population-level maternally-derived DENV PRNT titers, corresponding to the y-axis on the right hand side. Coloured points and lines show the median and 95% credible interval estimates of the expected age distribution of hospitalised infant dengue cases estimated using the different cohort-assay reconstructed datasets. **(C)** The mean age and 95% confidence intervals of infant (≤12 months of age) hospitalised dengue cases by year from 1974 to 2014 are shown by the grey points and lines. The blue line and ribbon show the mean and 95% confidence interval estimates from a linear regression model. **(D-F)** Estimated titer distributions associated with the age of increased risk of hospitalised dengue disease in infants. Estimates from the Bangkok PRNT, Bangkok HI and Kamphaeng Phet HI titers are shown in Panels D-F respectively. The grey dashed line and shaded ribbon show the median and 95% credible interval estimates of the probability of dengue hospitalisation by HI titer in the wider KPP cohort, estimated in Salje et al., corresponding to the y-axis on the right hand side of the panel (6).

We combined the reconstructed maternally-derived titers with the age distribution of hospitalised infant DENV cases, to quantify the relationship between infant titers and the risk of dengue hospitalisation. This approach assumes that antibody levels at the time of infection are the sole determinant of hospitalisation risk, with the relationship between antibody titer and enhanced risk of hospitalisation given infection following a log-normal distribution. We also assume that the distribution of maternally-derived antibody titers observed in our cohorts are representative of infants in the wider population in Thailand. We find that assuming a log-normal distribution of pre-existing risk-associated DENV titers is able to accurately reconstruct the age distribution of hospitalised infant dengue cases (Figure 3B). We estimate that for PRNT_50_ titers in the Bangkok cohort, the risk of hospitalisation is highest for infected infants with titers of 6.0 (95%CrI: 5.7-6.6), shown in Figure 3D, with large variation such that 95% of the risk of hospitalisation occurs between PRNT_50_ titers of 4.2 and 4022.1. For HI titers in the Bangkok cohort, we estimate that peak risk occurs at HI titers of 3.2 (95%CrI: 3.0-3.4), with 95% of risk occurring between HI titers of 1.3 and 27.4 (Figure 3E). For HI titer data in the Kamphaeng Phet cohort, we estimate a peak risk at titers of 5.2 (95%CrI: 4.9-5.6) with 95% of risk occurring between HI titers of 2.1 and 217.4 (Figure 3F). We compare this to previous estimates of the probability of dengue hospitalisation by HI titer inferred for older children and adults from a related cohort in Kamphaeng Phet between 1998-2002, where the same HI assay was used (grey dashed line, Figure 3F) (6). We explored alternative functional forms relating antibody titer levels and risk of hospitalisation upon infection (Figure S5). We found that models that assumed a constant risk of hospitalisation irrespective of titer, and models that assumed risk was concentrated in titers below a threshold could not explain observed age distributions of hospitalised infant dengue cases.

Finally, we explore the potential effects of long-term changes in the epidemiology of dengue and population birth trends in Thailand on infant dengue age-specific risk profiles. A decline in the force of DENV infection over the last 40 years has led to an increasing mean age of reported dengue cases from 8.2 years in 1981 to 29.7 years in 2017 (Figure 4A). Over this same timeframe, fertility rates in Thailand have declined from an average of 3.4 children per woman in 1980 to 1.5 children in 2019 (Figure 4B). The mean age of mothers at childbirth, however, has remained relatively stable over this period, declining slightly from 28.3 in 1980 to 27.3 in 2019 (Figure 4C). Using age-specific fertility data and assuming a linear decline in DENV transmission intensity from 0.18 in 1980 to 0.06 in 2019 (14, 15) we simulate the expected trends in infant dengue risk profiles over this 40-year period. We show how, under these assumptions, the cumulative probability of experiencing a DENV infection by 12 months of age declined from 0.16 in 1980 to 0.06 in 2019, however, the mean age of infant infection increased only slightly, from 5.8 to 5.9 months over the same period (Figure 5D). We estimate a 26.7% reduction in the proportion of mothers who have experienced ≥2 DENV infections at the time of birth - from an estimated 91.1% of mothers in 1980 to 66.8% of mothers in 2019 (Figure S6). We estimate that this reduction in lifetime DENV infections experienced by women giving birth translates to a decrease in population mean maternal DENV titers from 94.1 (95%CI: 92.6-95.6) in 1980 to 76.0 (95%CI: 73.9-78.1) in 2019.

**Figure 4.**
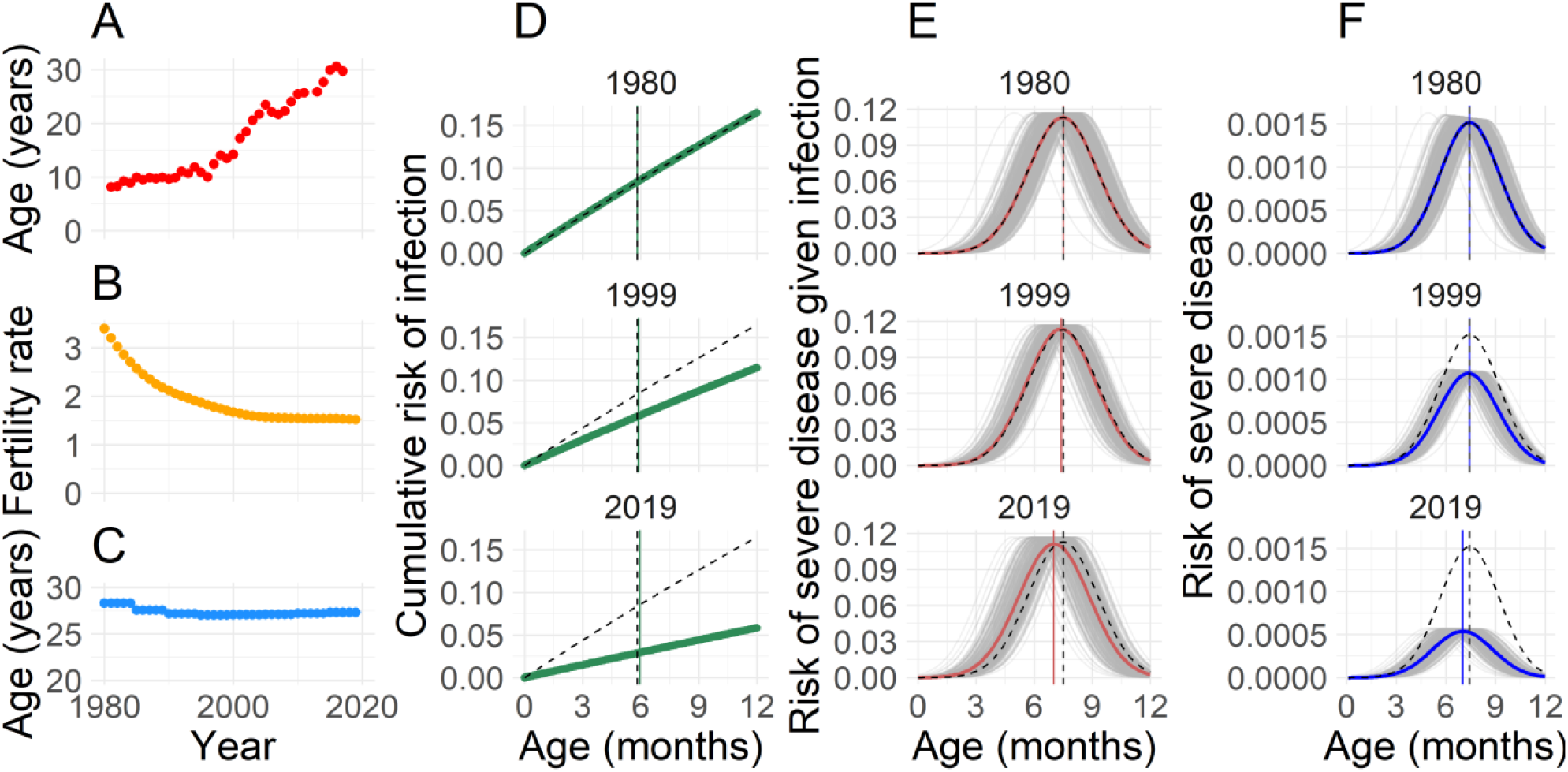
Impact of the shifting epidemiology of dengue on infant risk profiles. **(A)** Mean age in years of reported hospitalised dengue cases in Bangkok from 1981-2017. **(B)** Average number of live births per woman by year in Thailand from 1980-2019 (28). **(C)** Mean age of childbearing in Thailand by year from 1980-2019 (28). **(D)** Cumulative risk of infant DENV infection in the years 1980, 1999 and 2019 is shown by the green lines, while the dashed line indicates the cumulative risk of infection in 1980 as a reference point. The green vertical lines show the mean age of infant infection (<12 months old) by year and the dashed vertical lines show the mean age of infant infection in 1980. **(E)** Grey lines show the simulated risk of severe disease given infection for each individual infant by month of age in the years 1980,1999 and 2019. Red curved lines indicate the population mean risk of severe disease given infection by year, while the black dashed curved lines show the population mean risk of severe disease given infection in 1980, as a reference. Vertical red lines indicate the age of peak risk by year and vertical black dashed lines show the age of peak risk in 1980. **(F)** Grey lines show the simulated risk of severe disease for each individual infant by month of age in the years 1980,1999 and 2019. Blue curved lines indicate the population mean risk of severe disease, while the black dashed curved lines show the population mean risk of severe disease in 1980, as a reference. Vertical blue lines indicate the age of peak risk by year and vertical black dashed lines show the age of peak risk in 1980.

Using this same simulation framework we explore how this trend of declining population maternal DENV titers may impact infant DENV risk profiles in the first year of life. We estimate that decreasing mean DENV titers in birthing women reduces the mean age at which infants experience their highest titer-related risk of hospitalised dengue disease given infection, from 7.5 months in 1980 to 7.0 months in 2019 (Figure 5E, Figure S7). Subsequently, the mean age of infant risk of dengue hospitalisation is estimated to decline slightly from 7.4 in 1980 to 7.0 in 2019, with an expected 64.0% reduction in absolute risk of hospitalised infant dengue disease (Figure 4F). This is consistent with reported dengue case data over this same time period where the proportion of all dengue cases in Bangkok attributed to infants <1 year of age has declined significantly from 3.2% in 1981 to 0.6% in 2017 (Figure S8).

## Discussion

We developed mathematical models to investigate the relationship between infant maternally-derived DENV antibody titers and the associated risks of hospitalised dengue disease, using detailed longitudinal serological data and dengue hospitalisation data. By probabilistically reconstructing maternally-derived antibody titer distributions that are associated with the age of increased risk of hospitalisation, our analysis provides an improved understanding of the mechanistic relationship between antibody concentration and risk of hospitalised dengue disease in infants, in addition to identifying quantitative markers of infant dengue risk which can be used to guide public health action.

Our estimated distribution of HI titer concentrations that are associated with an enhanced risk of infant hospitalisation in the Kamphaeng Phet cohort are similar to the HI titers estimated to enhance the risk of hospitalisation among the wider population experiencing a post-primary infection (peak risk occurring at titers of approximately 5 in infants versus 12 in older individuals) (6). This adds evidence for the common mechanism, ADE, placing both infants and older individuals at increased risk of severe DENV disease. Slight differences between the two estimated distributions could potentially be caused by differences in the antigenic properties of the circulating viruses in the two cohorts, which occurred at different time periods (2015-2021 and 1998-2003) (6, 16, 17), or potentially by qualitative differences in immune responses. We note that unlike older individuals experiencing post-primary infections, infants lack pre-existing B cell or T cell immunity, suggesting that antibody levels are likely the predominant driver of individual-level risk of severe dengue disease. While PRNTs measure neutralisation, HI assays measure antibodies that bind to virions and inhibit hemagglutination but are not necessarily neutralising. However, we observe good correlation between PRNT_50_ and HI titer values with a Pearson correlation coefficient, R, of 0.62 (Figure S9) and find largely consistent titer-associated risk windows inferred independently from the PRNT_50_ and HI data. We also observed a trend of increasing HI:PRNT ratio with increasing infant age in the Bangkok cohort, though further work is needed to understand the drivers of this trend (Figure S10).

A steady decline in DENV force of infection in Thailand over the last 40 years has been well documented (14, 15, 18), leading to an increasing mean age of dengue cases. This trend has been driven, at least in part, by simultaneously decreasing fertility rates, which has led to reduced numbers of susceptible individuals in the population (14). Here we demonstrate how reduced transmission intensity of DENV over time has likely also shifted the immune landscape such that women of childbearing age have experienced fewer DENV infections than in previous decades, leading to reduced levels of transplacentally-transferred DENV antibodies in newborns. We show how this has had a negligible impact on the mean age of infant hospitalised cases, despite a substantial reduction in maternally-derived DENV antibodies. In terms of the absolute risk of disease, however, we find that reduced transmission intensity, coupled with declining fertility rates has driven a large reduction in the absolute risk of hospitalised dengue disease posed to infants.

Despite the reduction in infant disease, it remains critical to protect infants during periods that they are most vulnerable to severe disease. Our estimates of the relative risk of hospitalised infant dengue disease by month of age (Table S1) can inform guidance as to when infants should be maximally shielded from infection, for example, through limiting exposure to mosquitoes by placing screens on doors/windows and/or the use of long-sleeved clothing (19, 20). Further, careful consideration should be given to the risk profile of young infants when planning the implementation of any future DENV vaccines. The potential long-term effects of DENV vaccines on infant risk of severe disease, through both changes in mothers’ antibody levels and changes in transmission intensity should be assessed, particularly in the case where vaccines induce lower titers than natural infection (21).

Our analysis highlights the valuable role that mathematical models can play in scenarios of poorly observed processes. Our results show there are substantial discrepancies between measured and underlying true titer concentrations from discretized measurement processes and lower limits of assay detection. Accounting for these measurement processes, we were able to reconstruct the dynamics of maternally-derived antibody titers in infants. We find that constant exponential decay models are able to capture the dynamics of maternally-derived dengue antibodies (Figure 2A-C and Figure S11) and our estimates of antibody half-life, ranging from a median of 29-47 days across cohorts and assays, are consistent with previous estimates. Previous studies investigating the persistence of maternally-derived dengue antibodies in Thai infants estimated a DENV antibody half-life of 41 days (22) serotype-specific antibody half-lives ranging from 33-53 days (23) while another study in Vietnamese infants estimated a half-life of 42 days (12). The use of parametric models that characterise antibody titer dynamics are especially useful in the context of assay limits of detection. Here, we use our estimates of antibody half-life to estimate titer values below the limit of detection of 1:10 and infer the associated risks of hospitalised dengue disease at all titer levels.

The cohort studies used in this analysis were not designed to investigate individual-level infant risk of severe dengue disease. Doing so would require very large cohort sizes in order to observe a substantial number of severe outcomes. Our estimates of the titer distributions associated with increased risk of hospitalised dengue are therefore derived from population-level probabilistic inferences rather than individual-level associations, assuming that the infants recruited to the cohort studies are representative of the wider infant population. Our analysis assumes maternally-derived DENV antibodies to be the sole driver of hospitalised dengue disease risk and that age has no independent effect. We also assume the age distribution of infants aged <1 year to be uniform by month. Despite these simplifying assumptions, our findings are consistent with a study of 13 Thai infants with dengue haemorrhagic fever or dengue shock syndrome that were estimated to have PRNT_50_ titers <10 at the time of infection (11). Our study highlights the need for additional focused mother-infant cohort studies with more frequent antibody measurements as well as active surveillance of DENV infections and associated illness occurring in the first year of life.

This study also highlights the complications caused by the non-standardization of antibody assays, where differences in the estimated titer windows of risk across cohorts/locations may be due to differences in the antigenic properties of the specific viruses included in each assay rather than due to fundamental differences in risk by serotype-specific titers (24, 25). This underscores the complexity of comparing inferences across locations and studies where both differences in the circulating lineages and differences in reference viruses used in the assay can impede direct comparisons of results. Importantly, our comparison of infant maternally-derived antibodies and antibodies in older individuals come from cohorts in the same region of Kamphaeng Phet, Thailand, and were estimated using data from the same HI assay run in the same laboratory (AFRIMS, Thailand) (6, 17). We note that infant cases caused by DENV-4 had a lower mean age compared to cases caused by other serotypes, and that DENV-4 also had the lowest neutralisation titers (but highest HI titers). However, given the reliance of the titer on the virus used in the assay, we cannot draw any mechanistic conclusions with these data alone.

Our analysis has allowed an increased understanding into the dynamics of infant dengue disease risk including the potential epidemiological and demographic drivers that shape these risk profiles. By probabilistically reconstructing the distribution of dengue antibody titer concentrations that place infants at enhanced risk of hospitalised disease given infection, we have provided an estimate of antibody-based correlates of risk that can inform control and prevention efforts. In the future, direct measurements of antibody levels could potentially be used for the identification of infants at risk of severe disease upon infection. As age-specific risks of infant dengue disease depend on local DENV transmission and immune dynamics, the characterisation and monitoring of local maternal DENV immune profiles in endemic countries will be crucial for informing maximal preventative and control measures.

## Methods

### Cohort data

We use DENV serological data from two longitudinal cohort studies in Thailand. The first study was an infant cohort study conducted in Bangkok from 2000-2003 (26). A total of 219 mother-infant pairs attending the Rajavithi hospital in Bangkok between November 2000 and March 2001 were enrolled in the cohort (26, 27). Mother and cord blood samples were collected at the time of birth. Of these 219 infants, 138 (63%), 116 (53%), 118 (54%), and 115 (53%) infants were followed up at 3, 6, 9, 12 months of age, respectively, where blood was drawn for serological testing. All blood samples were tested using DENV serotype-specific hemagglutination inhibition (HI) and 50% plaque reduction neutralisation test (PRNT_50_) assays (26). PRNT_50_ titers were measured on a continuous scale with a lower limit of detection of 10, while HI titers were measured on a discretized scale using 2-fold serial dilutions with a lower limit of 1:10. Mothers with a history of immune deficiency, received immunosuppressive treatment in the previous month or received blood in the previous 3 months were excluded from study participation. Premature infants, twins, or infants with severe congenital anomalies were also excluded from the study (26).

The second cohort study is an ongoing family cohort in Kamphaeng Phet, Thailand (17). Mother-infant pairs enrolled between September 2015 and July 2021 were included in our analysis. Blood was drawn from each mother prior to giving birth as well cord blood at time of birth. Infants were followed up at approximately 1 year of age where blood samples were taken. Additional blood samples were collected from a subset of infants in the first year of life as part of household-illness investigations within the cohort. DENV antibody titers were tested using serotype-specific haemagglutination inhibition measured on a discretized scale using 2-fold serial dilutions with a lower limit of detection of 1:10.

To account for possible infection-related increases in antibody titers, the time at which any infant’s mean titer (averaged across serotypes) increased was marked as a potential DENV infection. These and any subsequent samples of the same infant were then excluded from the analysis. Samples with measured concentrations below the lower limit of assay detection (1:10) were assigned a value of 5 for the calculation of geometric mean titers by age. Infants who provided at least three samples within the first 13 months and who had no evidence of a DENV infection were included in the final analysis. A total of 165 infants were included in the final analysis, with 42 infants from the Kamphaeng Phet cohort and 123 infants from the Bangkok cohort.

### Dengue hospitalisation data

To explore the age-specific risks of hospitalised infant dengue disease we used reported hospitalised dengue case data from Queen Sirikit National Institute of Child Health (QSNICH), located in Ratchathewi, Bangkok, reported between 1974 and 2014, as well as from Kamphaeng Phet hospital, located in Kamphaeng Phet province, between 1994 and 2019. The age, in months, of infants hospitalised with a dengue diagnosis was available from both hospitals. Both hospitals are large, tertiary care facilities that accept patients from throughout their respective provinces.

### Modelling maternally-derived antibody decay

We model the decay of maternally-derived DENV antibody titers from birth until 13 months of age. To account for the discretized measurement process of the haemagglutination inhibition (HI) assay as well as the lower limit of detection truncation of both the HI and PRNT assays, we consider “true” unobserved continuous-scale antibody titers as distinct but related quantities to the measured antibody titers. Two-fold dilutions of 1:10, 20, 40, 80, 160, and so on were used for HI assay measurements. As HI antibody concentrations are reported as the lower bound of the dilution (e.g. antibody concentrations in the range of 1:80 - 1:159 will be reported as 1:80) we consider the “true” titer, *A*_*i,s,t*_, for an individual, *i*, to serotype, *s*, at time, *t*, on a continuous scale that underlies the discretized observed titer, 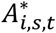, measured by the assay (6, 21). Analysis of titers was conducted on an adjusted *log*_2_ scale; 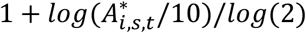, such that a measured titer of 1:10 = 1, 1:20 = 2, 1:40 = 3 and so on. We define the probability of the observed titer, given the true titer as shown in equation 1, where *f*(*u*) is a probability density function for a cumulative normal distribution, with estimated standard deviation *σ*.

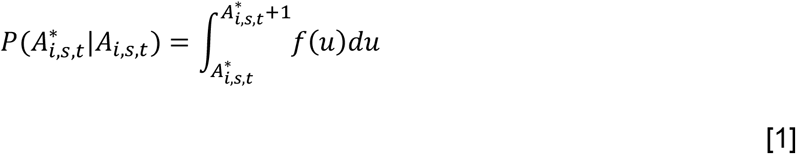

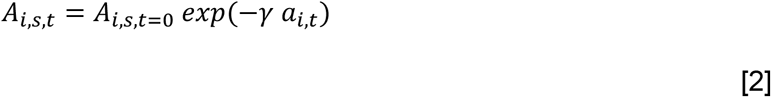

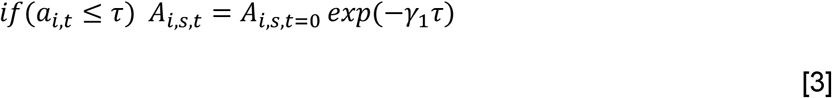

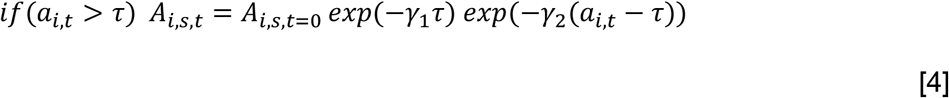

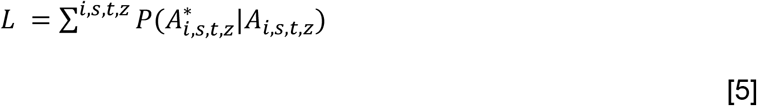

The initial “true” (unobserved) antibody titer of each infant was estimated and the “true” titers of each subsequent sample (including those below the lower limit of detection) were inferred on a continuous scale assuming either constant or biphasic rates of decay from this initial titer. We assumed identical decay dynamics across the 4 serotypes. Where a constant rate of decay was assumed, “true” antibody titers were calculated as shown in equation 2, where γ is the estimated rate of decay and *a*_*i,t*_ is the age of infant *i* at time *t*. For biphasic decay dynamics, the age at which the rate of decay changes, *τ*, was additionally estimated, and “true” titers were estimated as shown in equations 3–4. The full model likelihood was evaluated by summing the probability of observed titers given the estimated “true” titers across individuals *i*, serotypes *s*, time points *t*, and assay-cohorts *z*, as shown in equation 5.

We consider both a scenario where rates of antibody decay are assumed to be the same across different assays and cohorts (designated as an “ensemble” model) and a scenario where we allow independent rates of decay for each assay and cohort (designated as an “assay-cohort model”). Subsequently, 4 independent model parameterizations were considered; an ensemble constant decay, an ensemble biphasic decay, an assay-cohort constant decay and an assay-cohort biphasic decay model. The deviance information criterion (DIC) was calculated for each model specification and used to compare model performance. We fit exclusively to infant samples taken between birth (cord blood) and 13 months of age.

All model fitting was conducted in CmdStanR version 0.3.0. Each model was run with 3 chains of 5,000 iterations each, with a burn-in period of 1,000 iterations.

### Reconstructing the age distribution of hospitalised infant dengue cases

We combine the estimated “true” continuous-scale antibody titer model results and the observed age distribution of hospitalised infant DENV cases to infer the distribution of antibody titers that are associated with the age of enhanced risk of hospitalised dengue disease given infection. We assume that dengue antibody titer measurements within the two cohorts are representative of the wider populations and that maternally-derived dengue antibody titers at the time of DENV infection are the sole determinant of risk of hospitalised disease. Additionally assuming these sub-neutralising, infection-enhancing titers to follow a log-normal distribution with mean, *μ*, and standard deviation, *ε*, the probability density of maternally-derived dengue antibody titers falling within this distribution of risk-associated titer levels is calculated for each infant, *i*, by age, *a, ρ*_*i,a*_, between 0 and 12 months. The expected number of hospitalised infant dengue cases at age *a* (in months), Ĉ_*a*_, is calculated as the sum of individual-level probabilities at each month of age, multiplied by the probability of infection given a monthly average transmission intensity, *λ*, and reporting coefficient, *β*, which incorporates both the underlying population size and the probability of being a reported hospitalised dengue case. Dengue transmission intensity, *λ*, is assumed to be endemic and constant in time.

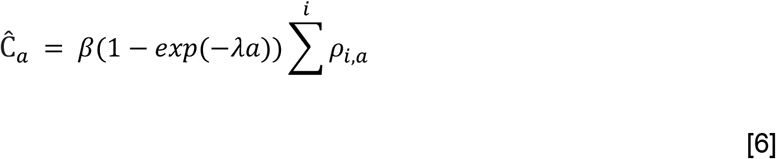

Age-specific counts of hospitalised infant dengue cases were assumed to follow a poisson distribution. The likelihood of this model is calculated as shown in equation 7. Model parameters were estimated independently for each cohort and assay combination. Model fitting was conducted in CmdStanR version 0.3.0. Each model was run with 3 chains of 5,000 iterations each, with a burn-in period of 1,000 iterations.

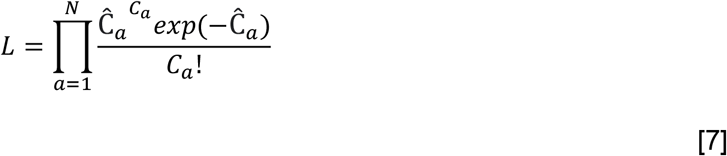

### Simulation testing of titer-related infection-enhancing mechanisms

To explore the possible maternally-derived antibody titer-related mechanisms that may enhance dengue severity, we simulate the expected age distribution of hospitalised infant dengue infections via a number of mechanisms. We consider 4 scenarios of the potential relationship between infant maternally-derived antibody titers and risk of hospitalised dengue disease upon infection (illustrated in Figure S4). We consider a baseline scenario, A, with a constant risk of hospitalised infant dengue disease upon infection, irrespective of antibody titer levels. In scenario B, we consider a mechanism whereby infants with antibody titers below a certain threshold have a constant risk of hospitalised disease, given a DENV infection. Above this threshold, infants are completely protected from hospitalised disease. In scenario C we consider a constant risk of severe disease upon infection within a specific window of antibody titer levels. Antibody titers above and below this window of risk are assumed to be protected from hospitalised disease upon infection. In scenario D we consider a mechanism whereby the risk of hospitalised disease upon infection follows a log-normal distribution with a given mean infection-enhancing titer level and standard deviation. In all scenarios we assume a time-constant, endemic transmission intensity of 0.12 such that the risk of infection increases exponentially with increasing age. We use our estimates of “true” maternally-derived antibody titers in the population, *A*_*i,t*_, to calculate the probability density of each infant, *i*, at age, *a*, being within the assumed distribution of risk-associated titers, as well as the probability of experiencing an infection at each age. We calculate the mean risk of hospitalised disease across all infants by age to approximate the expected age distribution of hospitalised infant dengue disease under each of the assumed scenarios, as shown in equation 8. Here, *D*_*a*_ is the expected population mean risk of hospitalised disease at age *a, d*_*i,a*_ is the individual-level risk of hospitalised disease at age, *a*, for infant, *i*, and *N* is the total number of infants.

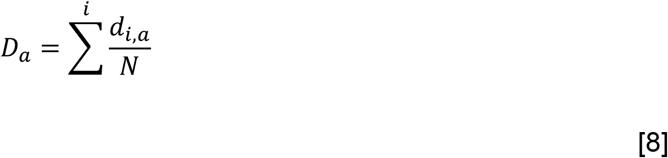

### *Modelling temporal trends in infant risk of* hospitalised *dengue disease*

We use data on age-specific fertility rates in Thailand, available in 5-year age groups between 15-49 years of age, for the years 1980-2019 from the United Nations World Population Prospects (28). We assume that the transmission intensity of DENV in Thailand declines linearly from 0.18 to 0.06 between 1980 and 2019, as estimated in previous analyses (14, 15), and that it was constant at 0.18 in years prior to 1980. Using the annual age-specific fertility rates, we calculate the number of mothers giving birth in 5 year age groups in the range of 15-49 years, for each year between 1980-2019. The age of each mother is drawn from a uniform distribution between the lower and upper bounds of their respective age groups. The probability of being susceptible, monotypic immune and multitypic immune to dengue is subsequently calculated for each mother as shown in equations 9-10.

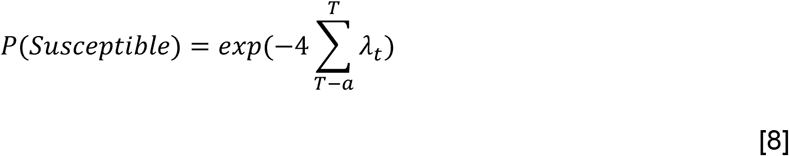

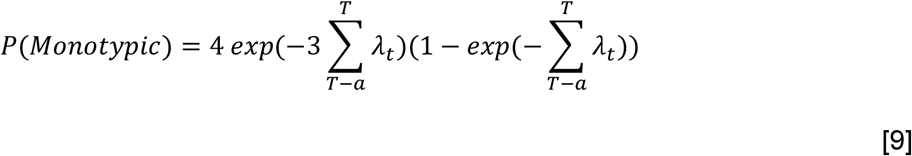

Here, *λ*_*t*_ is the DENV serotype-specific transmission intensity in year *t*, assuming all 4 serotypes to circulate with equal intensities. *T* is the year at time of giving birth and and *T* − *a* the year the mother was born, with *a* representing the age of the mother. The probability of having experienced ≥2 DENV infections by year *T* is simply calculated as 1 − *P*(*Susceptible*) − *P*(*Monotypic*). For each mother we calculate a weighted mean expected DENV titer at the time of giving birth. We assume that DENV antibody titers are drawn from normal distributions, truncated between 0 and infinity. We assume that DENV antibody titers of susceptible mothers have a mean of 5 and standard deviation 5, that monotypic titers have a mean of 40 and standard deviation of 10, and that multitypic titers have a mean of 100 and standard deviation of 20. As quantifying the specific titers linked to each serostatus is largely unknown and will differ by assay and cohort, we focus on the trends in titers in mothers and infants over time. We assign each birthing mother a mean DENV titer by drawing a random value from each of the three distributions and calculating the weighted mean titer, with weights equal to the respective probabilities of being susceptible, monotypic and multitypic to dengue. We assume this titer to be equal to the maternally-derived DENV antibody titer present in the respective infants and birth and model the decay of these titers in each infant assuming a constant exponential rate of decay (equation 2). Assuming the DENV titers associated with enhanced risk of hospitalised disease given infection follow a lognormal distribution, the joint probability of an infant experiencing a DENV infection and risk of hospitalised disease given titer levels was calculated by age for each infant.

## Data Availability

All code and anonymized data to reproduce the analysis will be made available.

## Ethics statement

The cohort study in Kamphaeng Phet was approved by Thailand Ministry of Public Health Ethical Research Committee, Siriraj Ethics Committee on Research Involving Human Subjects, Institutional Review Board for the Protection of Human Subjects State University of New York Upstate Medical University, and Walter Reed Army Institute of Research Institutional Review Board (protocol #2119). The Bangkok cohort study protocol was approved by the Ethics Committee of the Ministry of Public Health of Thailand. Our analysis is based on de-identified antibody titer data as well as de-identified hospital case data only.

Material has been reviewed by the Walter Reed Army Institute of Research. There is no objection to its presentation and/or publication. The opinions or assertions contained herein are the private views of the authors, and are not to be construed as official, or as reflecting true views of the Department of the Army or the Department of Defense. The investigators have adhered to the policies for protection of human subjects as prescribed in AR 70–25.

## Supplementary Information

**Figure S1.**
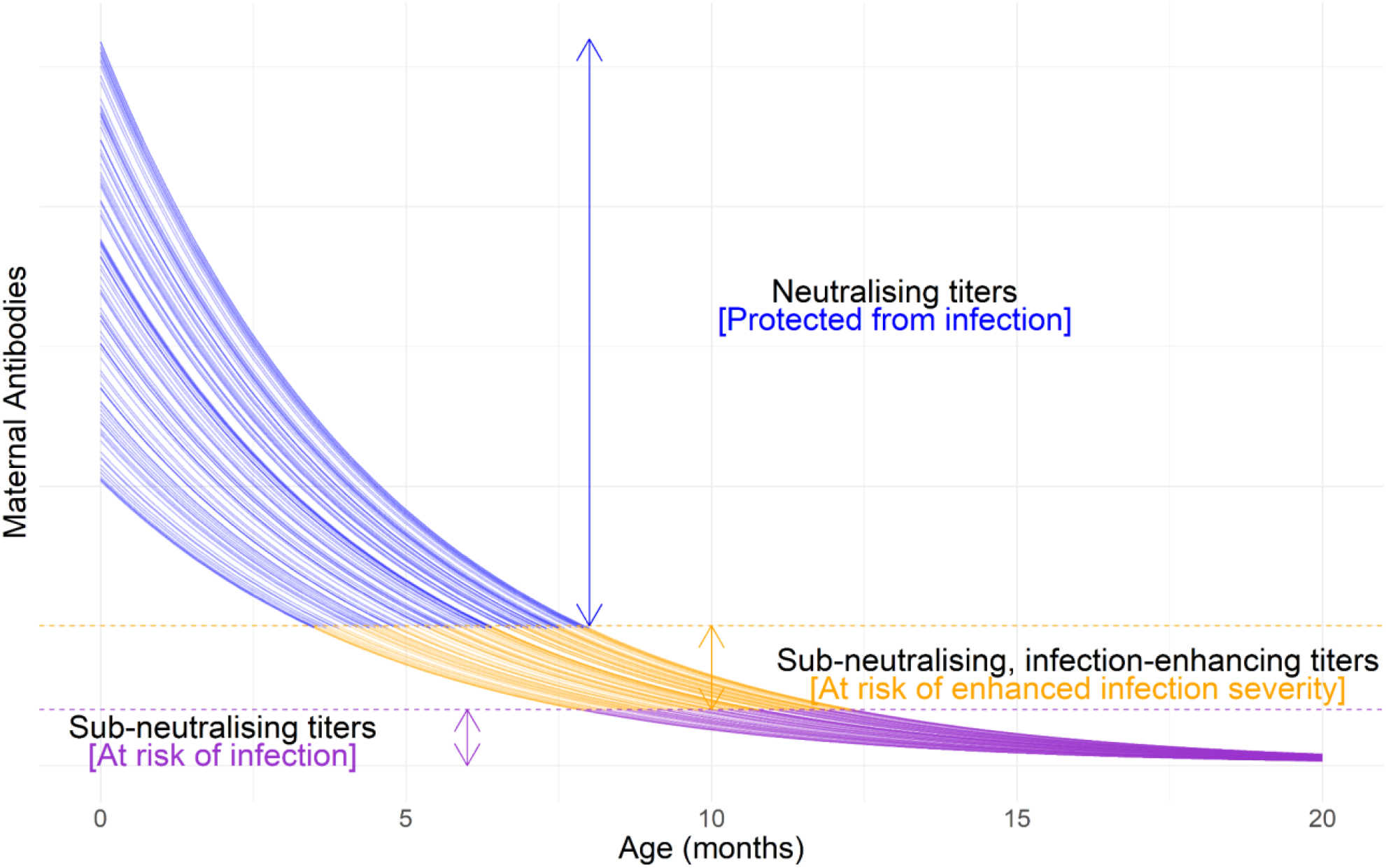
Illustration of maternally-derived DENV antibody decay dynamics and associated risks of infection and disease. Solid lines show the simulated maternal DENV antibody titers of 100 infants assuming an antibody half-life of 40 days. The colour of the lines indicate the expected risk profile of infants. Neutralising maternal antibody titers that protect infants against infection are shown in blue. Maternal antibody titers subsequently decay to levels that are sub-neutralising but infection-enhancing, shown in yellow, which place infants at risk of infection and at increased risk of severe disease, given infection. As maternal antibodies decay further, shown in purple, their ability to enhance heterologous infections is lost, reducing the risk of severe disease given infection, though infants will remain at risk of infection with these titer levels.

**Figure S2.**
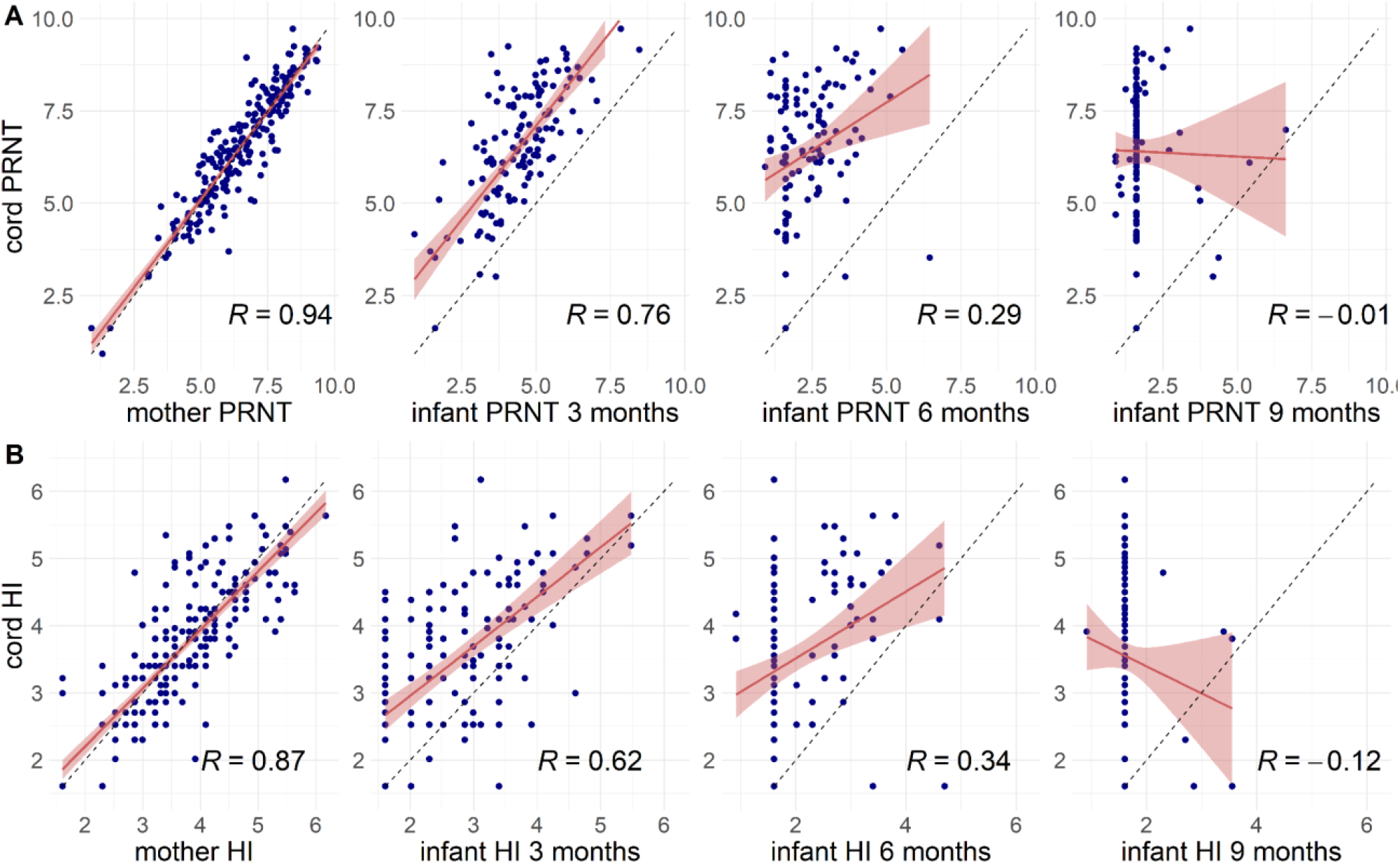
Correlation in cord titers at birth with mother’s titers and infant titers by age in the Bangkok cohort for both PRNT (A) and HI (B) assays. Blue points show individual PRNT and HI titer values on a log scale. The red line and ribbon show the mean and 95% confidence intervals of a linear regression model fitted to these data and corresponding Pearson correlation coefficients, R, are shown in black. The black dashed line indicates where log cord titers would be equal to mother or infant titers.

**Figure S3.**
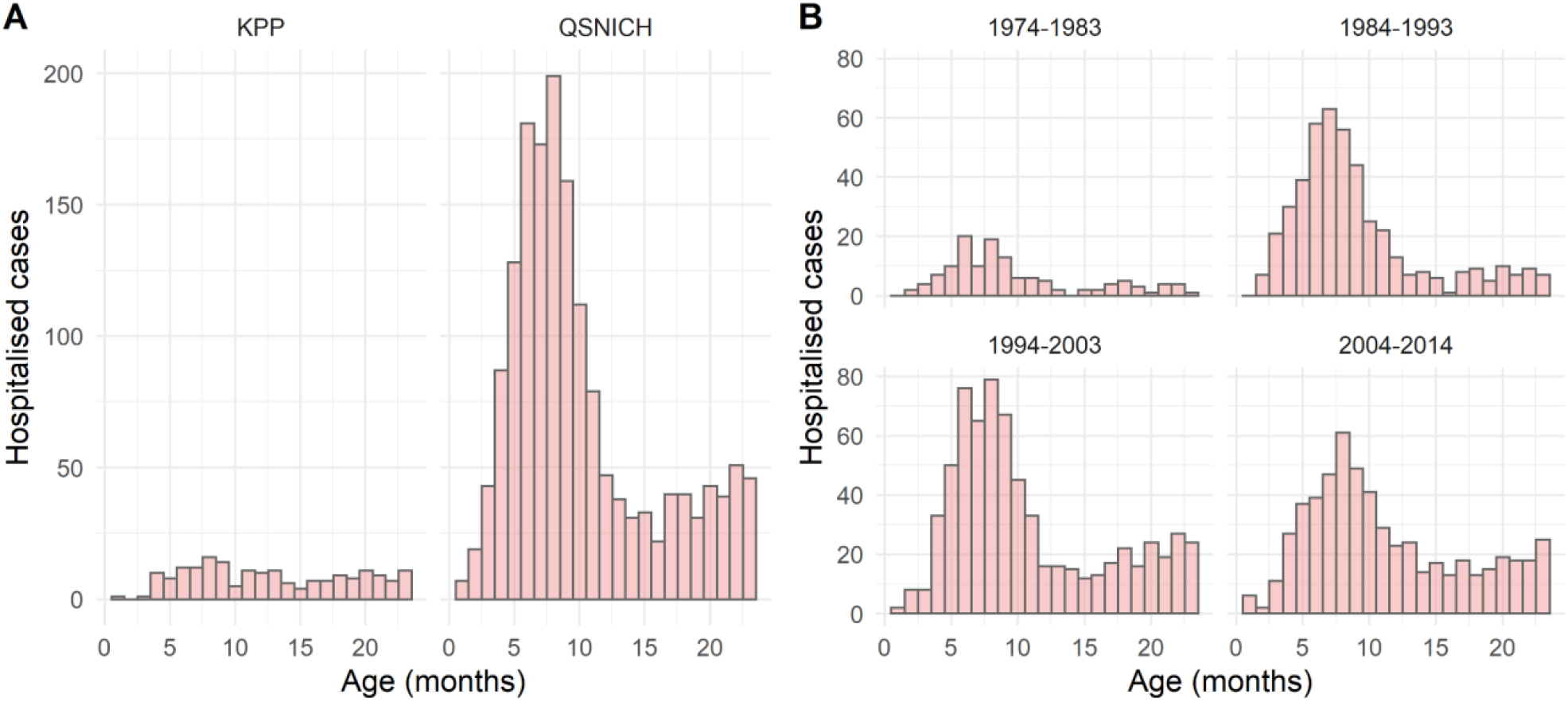
Age distribution of hospitalised infant dengue cases by hospital (A) and by decade (B). Data from Kamphaeng Phet (KPP) hospital was available for the period 1994-2019 and for the period 1974-2014 from Queen Sirikit National Institute of Child Health (QSNICH).

**Figure S4.**
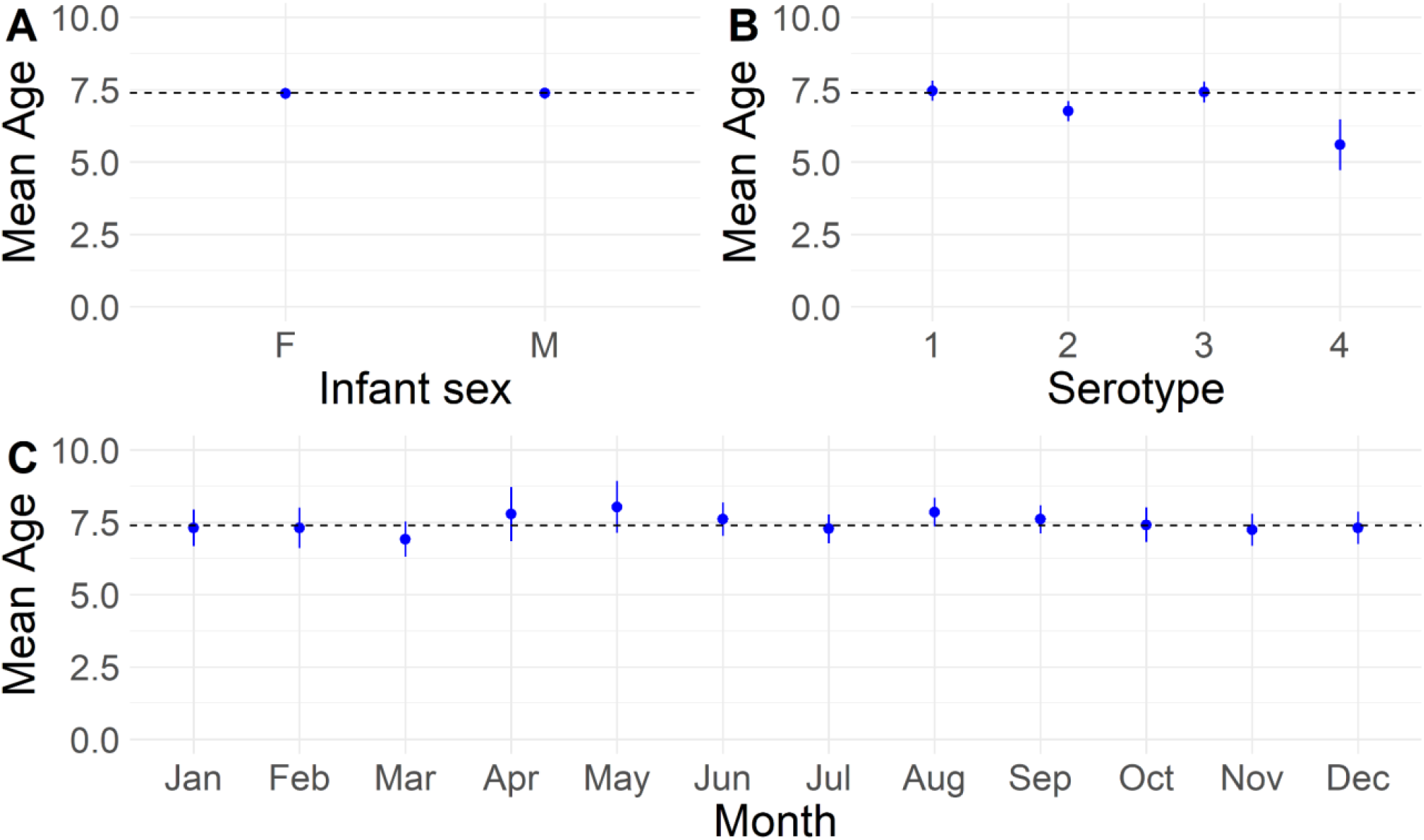
Mean age, in months, of hospitalised infant dengue cases by sex (A), infecting serotype (B) and by month of hospitalisation (C). Blue points and lines indicate the mean and 95% confidence interval. Black dashed lines indicate the overall mean age of infant hospitalised dengue cases.

**Figure S5.**
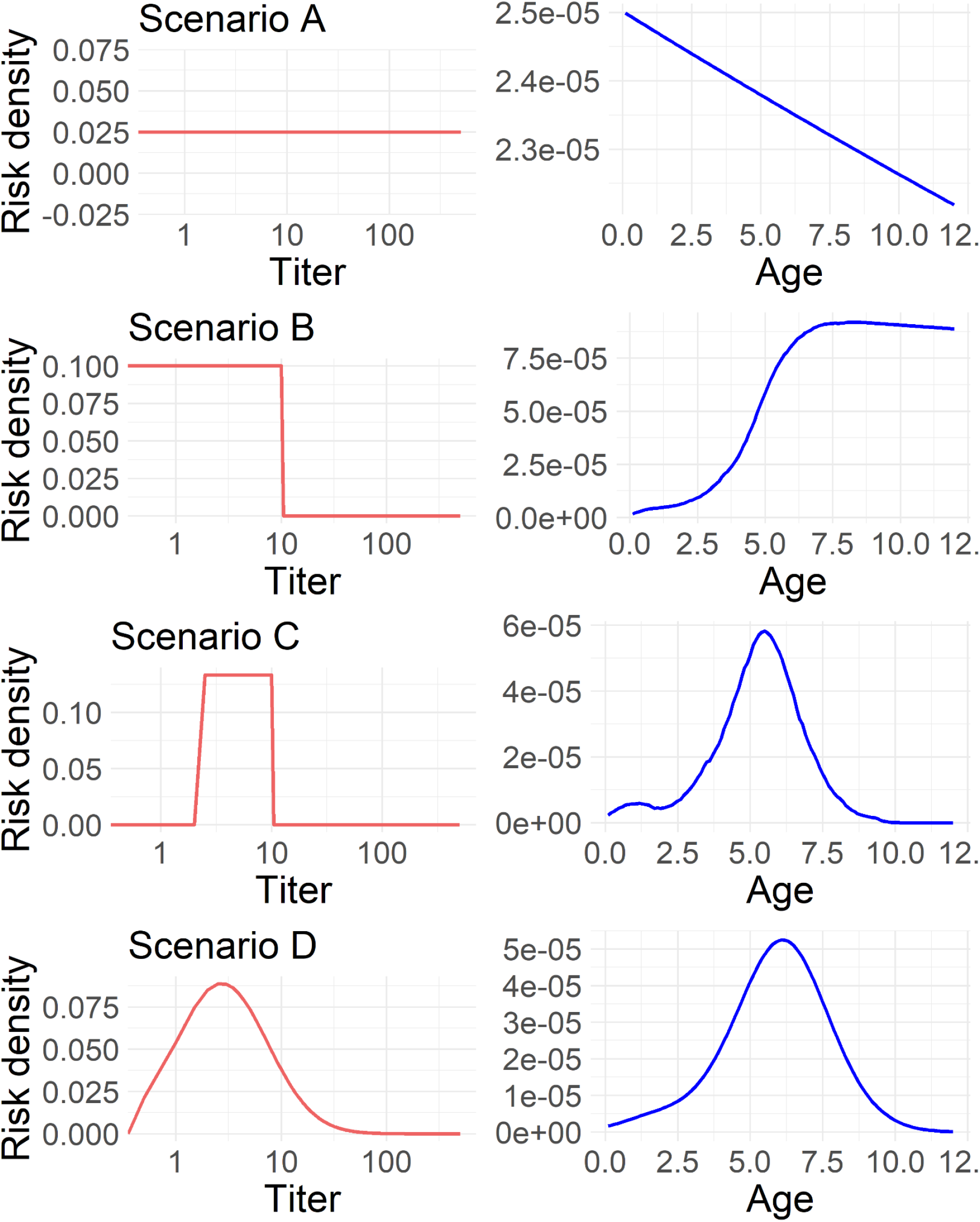
Simulated age distributions of severe infant dengue infections under four considered scenarios of titer-related risk. Red lines show the assumed relationship between maternally-derived anti-DENV antibodies and risk of severe dengue disease upon infection. Blue lines show the simulated age distributions, in months, of severe dengue disease resulting from each of the considered scenarios.

**Figure S6.**
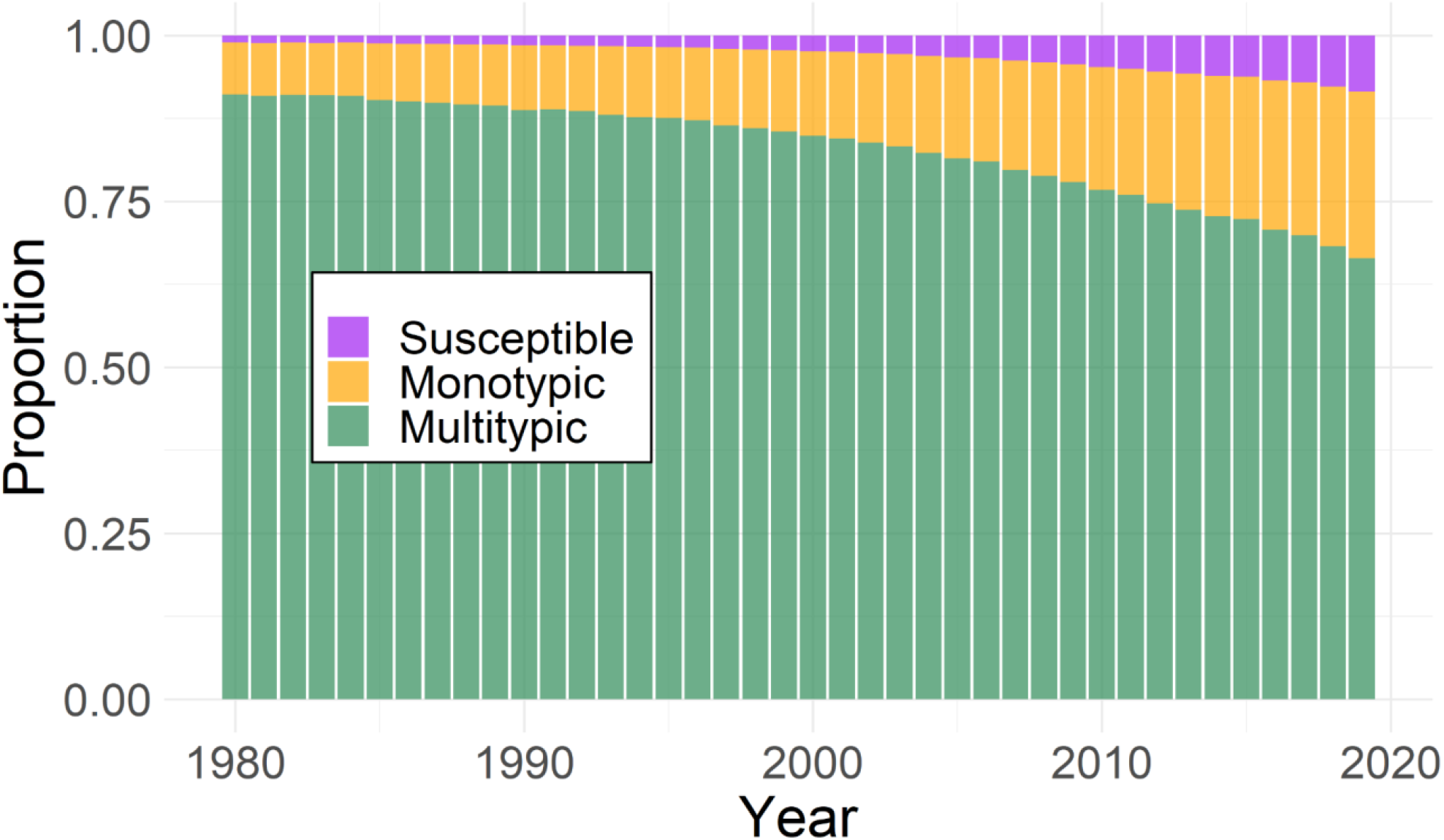
Estimated proportion of birthing mothers in Thailand that are susceptible, monotypic immune and multitypic immune to dengue virus by year, 1980-2019.

**Figure S7.**
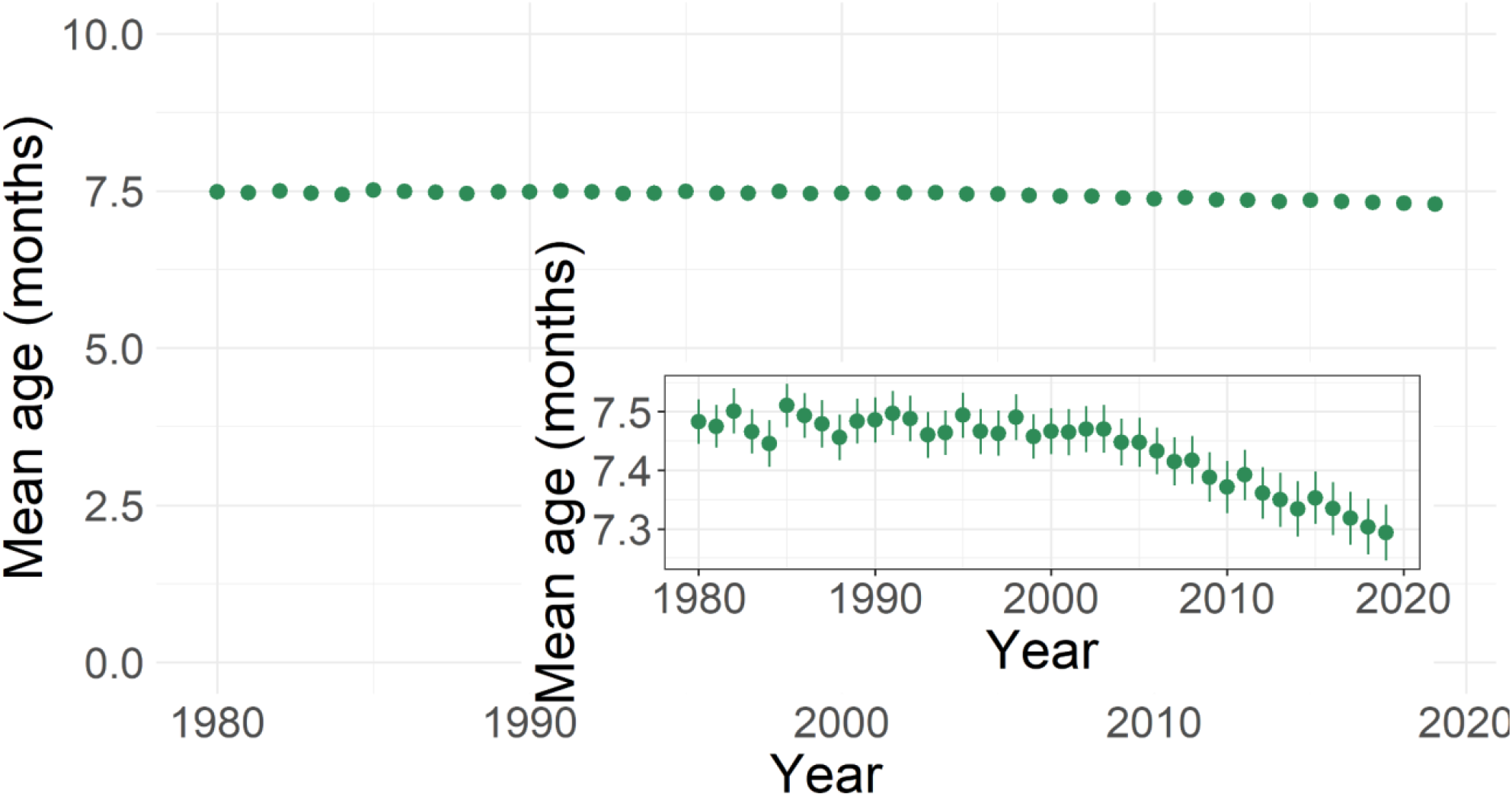
Mean age, in months, of hospitalised infant dengue cases estimated from simulation study. Green points and lines indicate the mean and 95% confidence intervals. The inset plot shows the same data on a zoomed y-axis.

**Figure S8.**
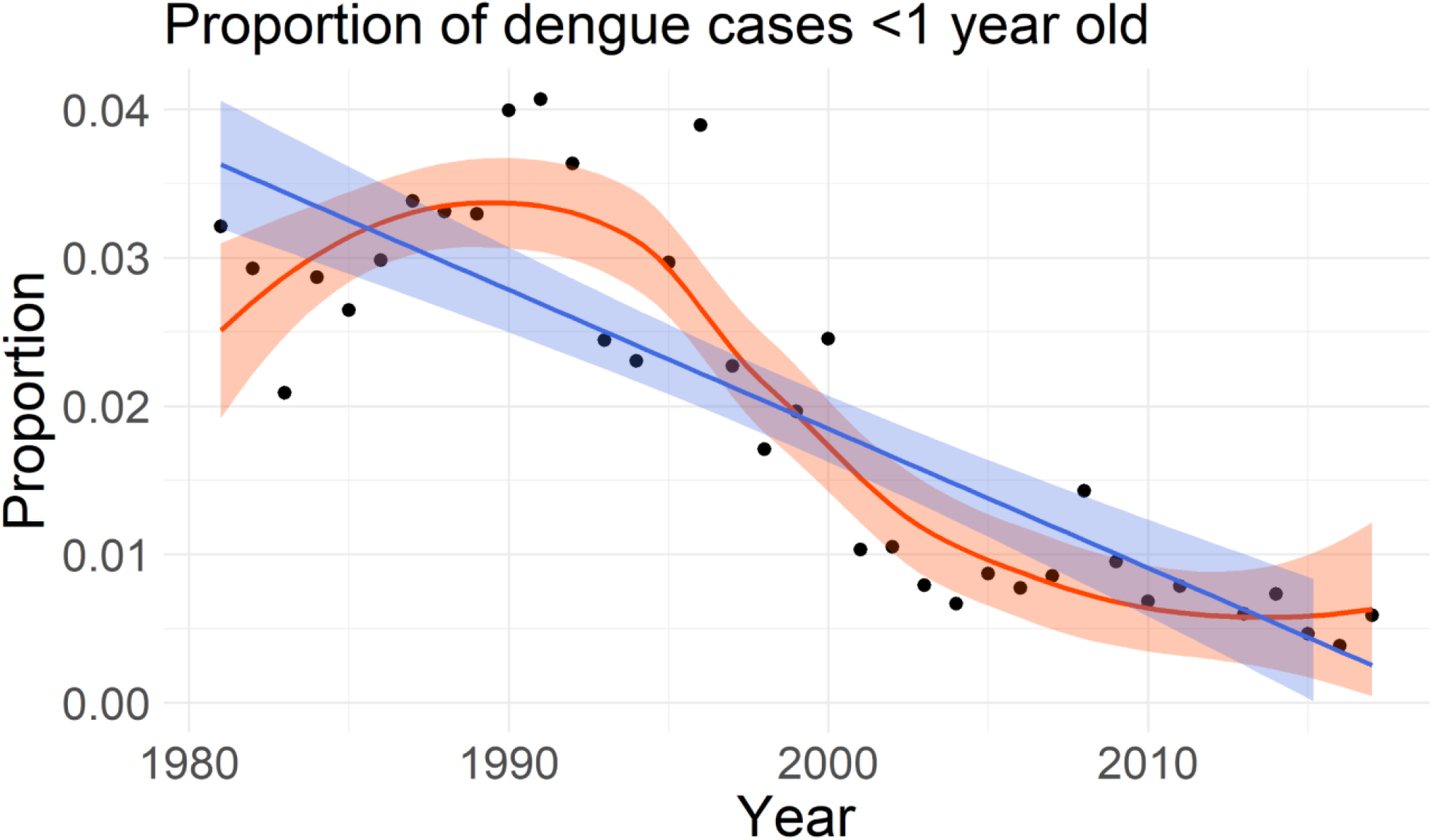
Annual proportions of all dengue hospitalisations reported in Bangkok that were attributed to infants <1 year of age between 1981 and 2017. Black points show the annual proportions of infant hospitalisations. The blue line and shaded ribbon show the mean and 95% confidence interval fit of a linear regression model, while the orange line and ribbon show the mean and 95% confidence interval fit of a loess model to the observed data.

**Figure S9.**
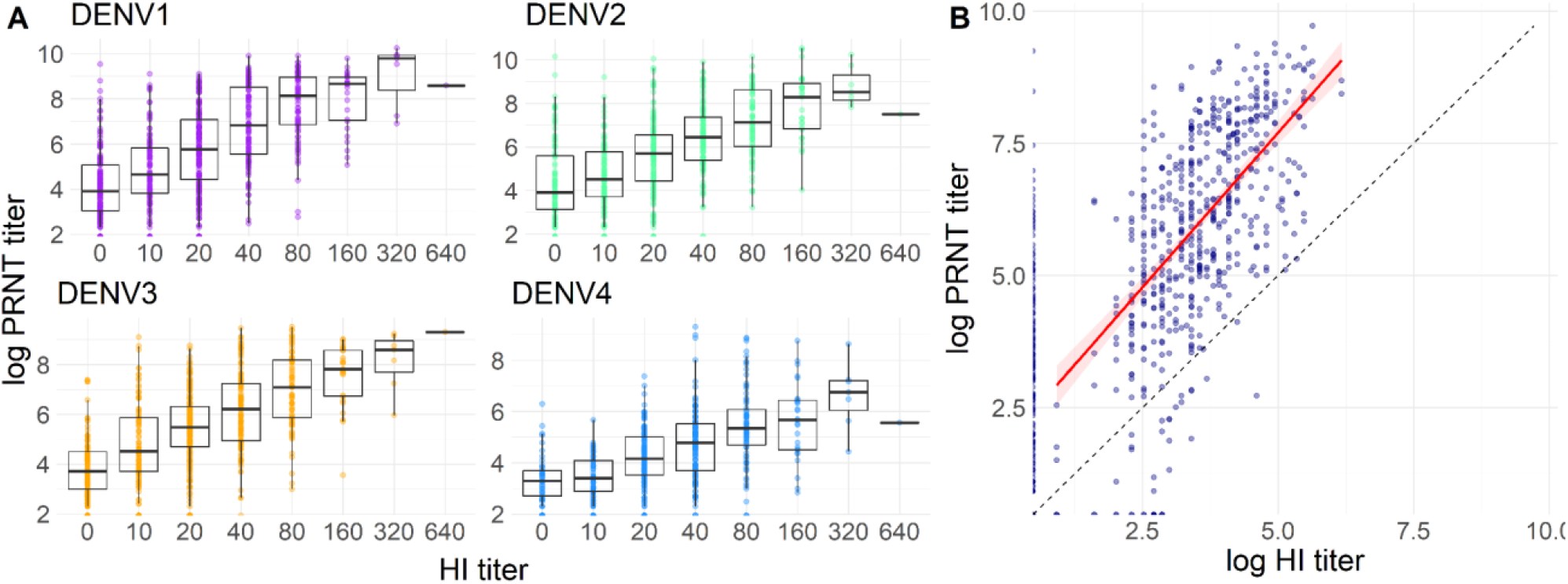
Correlations in PRNT and HI titers in the Bangkok cohort. (A) Coloured points show individual-level HI and log PRNT titers by serotype. Boxplots show the median and interquartile range of log PRNT titers per HI reading. (B) Blue points show individual-level log HI and PRNT titers averaged across serotypes, with the red line and ribbon showing the mean and 95% confidence interval linear regression model fitted to these data points (Pearson correlation coefficient=0.62 (95%CI: 0.58-0.65)). The black dashed line indicates where log HI values would be equal to log PRNT values.

**Figure S10.**
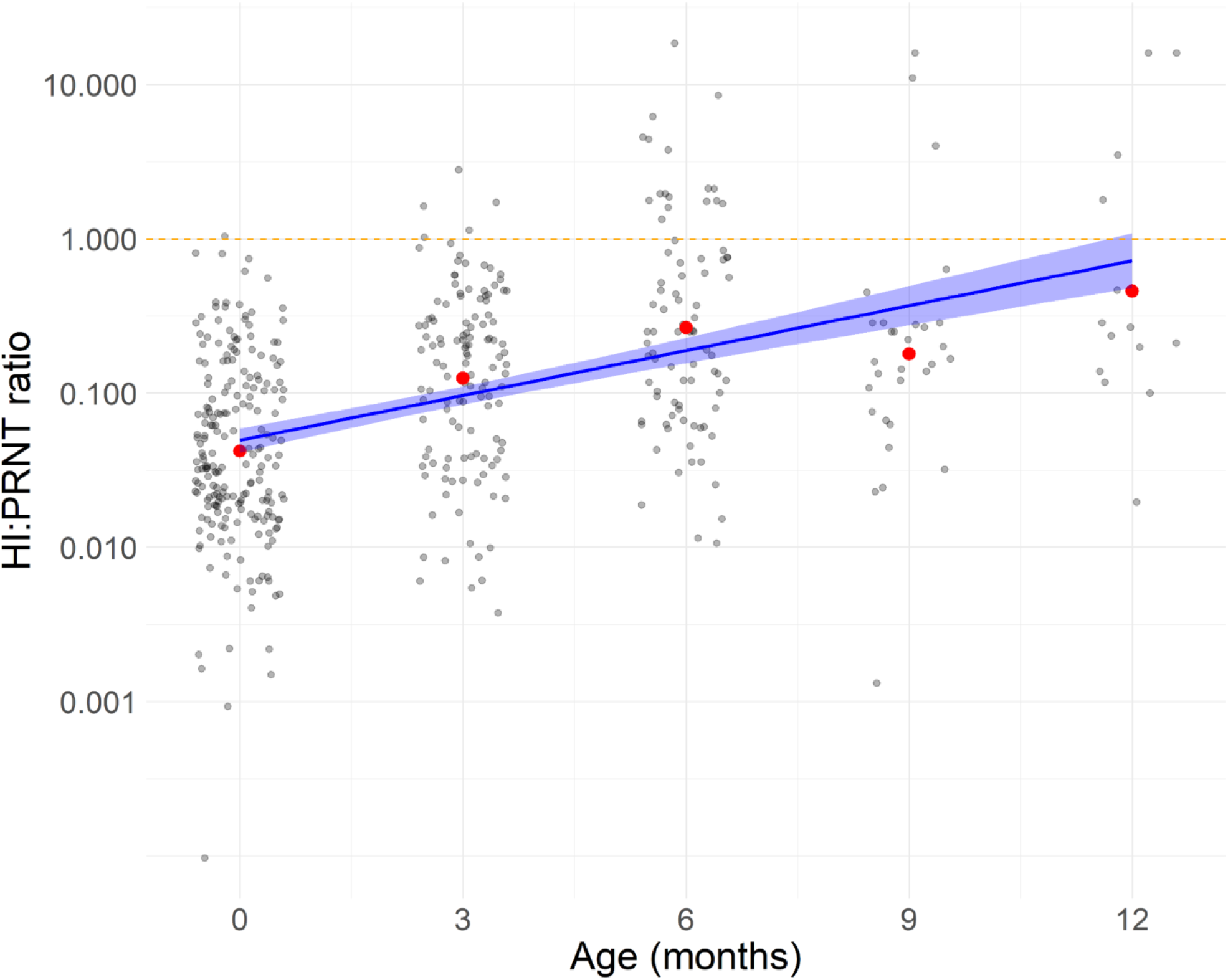
HI:PRNT ratios observed in the Bangkok cohort by month of age. Black points show individual infant HI:PRNT ratios and red points show the mean ratio by each age group. The blue line and ribbon show the mean and 95% confidence interval fit of a linear model to the observed ratios. The orange dashed line at 1 indicates where HI and PRNT titers become equivalent.

**Figure S11.**
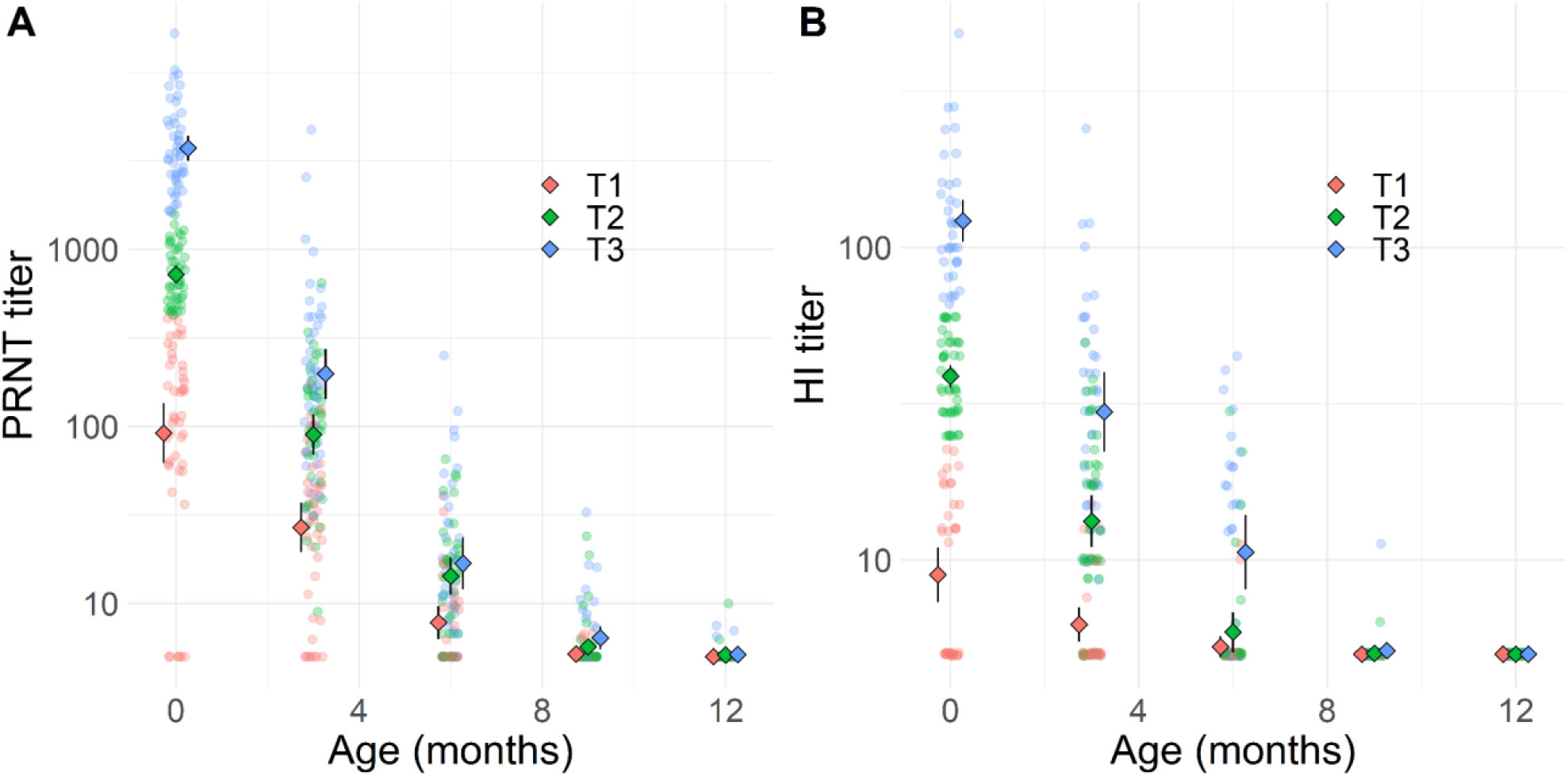
Geometric mean titers by age grouped by tertiles of cord blood titers at birth in the Bangkok cohort for PRNT (A) and HI (B) assays. Shaded points show individual DENV titers by age, averaged across the 4 serotypes, with colours indicating the cord blood tertile group of each individual. Coloured diamonds and black lines indicate the group-specific geometric mean titers by age and corresponding 95% confidence intervals.

**Table S1.** Antibody decay model comparison. Median and 95% credible interval estimates of antibody half-life are shown for each of the considered models. Deviance information criterion (DIC) was used to compare model performance.

| Model | Age (months) | Half-life in days (95%CrI) |  |  | DIC |
| --- | --- | --- | --- | --- | --- |
|  |  | KPP HI | Bangkok HI | Bangkok PRNT |  |
| Ensemble | 0-13 | 32.7 (31.8-33.6) | 32.7 (31.8-33.6) | 32.7 (31.8-33.6) | 5622.8 |
| Assay-Cohort | 0-13 | 38.6 (35.6-41.5) | 47.1 (44.9-49.3) | 28.5 (27.6-29.4) | 4975.8 |
| Ensemble Biphasic | 0-3 | 36.8 (34.3-51.4) | 36.8 (34.3-51.4) | 36.8 (34.3-51.4) | 6447.9 |
|  | 3-13 | 29.9 (27.7-31.4) | 29.9 (27.7-31.4) | 29.9 (27.7-31.4) |  |
| Assay-Cohort Biphasic | 0-5 | 38.5 (33.0-47.2) | 52.1 (48.2-67.3) | 28.6 (27.2-32.2) | 19599.3 |
|  | 5-13 | 38.7 (30.7-50.9) | 34.7 (27.2-43.9) | 28.4 (26.0-32.0) |  |

